# Effect of a new graphically modified Nutri-Score on the objective understanding of foods’ nutrient profile and ultra-processing – a randomised controlled trial

**DOI:** 10.1101/2022.11.18.22282494

**Authors:** Bernard Srour, Serge Hercberg, Pilar Galan, Carlos Monteiro, Fabien Szabo de Edelenyi, Laurent Bourhis, Morgane Fialon, Barthélémy Sarda, Nathalie Druesne-Pecollo, Younes Esseddik, Mélanie Deschasaux-Tanguy, Chantal Julia, Mathilde Touvier

## Abstract

**Context and objective:** When considering the health-related impact of foods, nutrient profile (content in salt, sugar, fibre, etc.) and (ultra-)processing are two complementary dimensions. The Nutri-Score, a summary graded front-of-pack label, already used in seven European countries, informs on the nutrient profile dimension, i.e. the one with the strongest evidence. Recently, mounting evidence linked ultra-processed food consumption to various adverse health outcomes, independently of their nutrient profile. To inform consumers about each of these two health-related dimensions of food (i.e., nutrient profile and ultra-processing), we aimed to test, in a randomised controlled trial, if a graphically modified version “Nutri-Score 2.0”, including a black “ultra-processed” banner, would improve the capacity of consumers to rank products according to their nutrient profile but also to detect those ultra-processed, compared to a no-label situation.

**Methods:** A total of 21,159 participants included in the NutriNet-Santé web-cohort were randomly assigned to a control arm (no front-of-pack label) or an experimental arm (Nutri-Score 2.0), and were presented an online interactive questionnaire with 3 sets of food products (8 cookies, 7 breakfast cereals, and 7 ready-to-eat meals) to rank according to their nutrient profile, and to identify ultra-processed foods. The primary outcome was the objective understanding of nutrient profile and ultra-processing, represented by a score of correct answers. Secondary outcomes were purchasing intentions and the healthiest-perceived product. Multinomial logistic regressions were performed.

**Results:** The Nutri-Score 2.0 increased significantly the objective understanding of both the nutrient profile dimension of food products from 0.9% to 24.2% (OR = 29.0 (23.4 – 35.9), p<0.001), and the ultra-processing dimension from 4.4% to 77.7% (OR = 174.3 (151.4 – 200.5), p<0.001), compared with no front-of-pack label. The results followed similar trends for cookies, breakfast cereals, and ready-to-eat meals. The Nutri-Score 2.0 also had a positive impact on purchasing intentions and on the products perceived as the healthiest, guiding consumers towards a better nutrient profile and non-ultra-processed products.

**Conclusion:** This randomised controlled trial demonstrates the interest of a front-of-pack label combining the Nutri-Score (informing on the nutrient profile dimension) with an additional graphic mention indicating when the food is ultra-processed, compared to a no-label situation. Our results show that participants were able to independently identify and understand these two complementary dimensions of foods.

**Trial registration number:** NCT05610930

## Introduction

Diet is recognized as one of the main modifiable risk factors for many chronic diseases, with 11 million deaths attributable to dietary factors in 2017 (1). Interpretative color-coded front-of-pack nutrition labels are considered as efficient tools to help consumers make healthier food purchases, and contribute to preventing nutrition-related diseases (2–5). They provide a quick and easy-to-use translation of the back-of-pack mandatory nutritional information and incentivize food manufacturers to improve the nutritional quality of their recipes (6–9).

In the framework of the European Farm to Fork strategy, the European Commission is expected to propose, by the end of 2022-early 2023, a harmonised mandatory front-of-pack nutrition labelling (10). The Nutri-Score is one of the options for this labelling regulation. It is a scientifically validated label reflecting the overall nutritional quality of food products (11) with 5 colours and letters, ranging from A-dark green to E-dark orange. Its algorithm is based on a modified version of the British Food Standards Agency nutrient profiling system (FSAm-NPS), originally aiming to regulate food advertising for children (12–14). This nutrient profiling system is computed on the basis of the composition per 100g of food (100 ml for beverage) in energy, sugars, saturated fats, sodium, dietary fibres, proteins as well as fruits, vegetables, legumes, nuts and rapeseed, walnut and olive oils. Convincing levels of evidence based on thousands of etiological studies have been established regarding the positive or negative impact of these nutritional components on the risk of chronic diseases (e.g., cardiovascular diseases, cancers, respiratory diseases) through different mechanisms (i.e., chronic inflammation, oxidative stress, gut microbiome, weight gain) (15): for instance, dietary fibres and fruits and vegetables play beneficial roles in the prevention of aerodigestive cancers (16), while diets high in saturated fat are associated with higher mortality from all-causes, cardiovascular disease, and cancer mortality (17). Furthermore, a diet of lower nutritional quality as reflected by FSAm-NPS has been associated with unfavourable health outcomes in several European cohorts, such as weight gain (18), asthma symptoms (19), cancers (20–22), metabolic syndrome (23), cardiovascular diseases (24,25) and mortality (26–29).

On the other hand, the past few years have seen a sharp increase in research on the health impact of foods by incorporating an additional key dimension: ultra-processing (30). In the past decades, scientists and international health organizations started developing classifications in order to categorize foods based on their level of processing, independently of their nutrient profile (31). One of the most extensively used classifications in cohort studies is the NOVA classification first developed in Brazil (32). The group with the highest level of processing according to NOVA, known as ultra-processed foods (UPFs). These products are generally submitted to intense physical and chemical processes, including hydrogenation, hydrolysis, extrusion, and pre-processing by frying, and contain food substances that are not usually found in domestic kitchens (such as maltodextrin, hydrogenated oils, modified starches), as well as flavouring agents, colorants, emulsifiers and other additives with “cosmetic functions”. Ultra-processed foods include for instance meat products made with mechanically separated meat, fish and chicken nuggets, instant noodles and dehydrated soups, chocolate and energy bars, carbonated drinks (sodas), vegetable patties (i.e. meat substitutes containing protein isolates and/or food additives), slimming products, powdered or fortified meal replacement shakes and snacks, and many other ready-to-consume food and drink products (33). In the last five years, associations between the consumption of UPFs and the risk of chronic diseases have been investigated in more than 50 cohort studies, conducted in various populations worldwide (e.g., Sun and Predimed cohorts in Spain, NutriNet-Santé in France, Nurses’ Health Study in the USA, UK Biobank) (34–37). These studies have identified direct associations with the risk of several chronic diseases, even after adjustment for components of the nutritional quality of the diet (sugar, salt, saturated fatty acids and energy), illustrating the complementarity of the two dimensions. Following these studies, several countries (e.g., Brazil, Israel, France) have included the objective of reducing UPF consumption as part of their dietary recommendations. While the two dimensions are related (i.e., on average, UPF tend to have a worse nutrient profile (38–49)), they are not collinear but rather complementary. Indeed, diet sodas with artificial sweeteners and food colourings do not necessarily exhibit a low nutritional value (since they contain no nutrients of concerns such as sugar or calories), yet they are definitely ultra-processed. Conversely, a 100% grape juice is not ultra-processed but yet have a low nutritional value overall (containing more than 160g of sugar/litre). Even though preliminary investigations are exploring new indices including both the nutrient profile and ultra-processing in the framework of exploratory research, current scientific knowledge does not allow combining these two dimensions to compute a single synthetic quantitative indicator able by itself to summarize the overall health value of food, used as a public health tool, due to a lack of scientifically validated weighting schemes for each dimension, (30). Thus, at this stage in research it appears important to provide consumers with tools that may simultaneously 1) inform them on the overall nutrient profile of the product and 2) help them identify which foods are ultra-processed.

A modified version of the Nutri-Score (Nutri-Score 2.0), containing an additional graphic mention when the product is ultra-processed, has been proposed (30). We aimed to study, in a randomised controlled trial design nested in the NutriNet-Santé cohort, the impact of this modified version on the objective understanding of foods’ nutrient profile, and on the identification of UPFs as primary outcomes. We also studied as secondary outcomes the impact of this label on purchasing intentions and the product perceived as the healthiest.

## Methods

### Trial design, participants, and general data collection

A two-arm parallel group randomised trial was conducted between April and June 2022. Participants were randomly recruited from the NutriNet-Santé cohort via an emailing campaign and were asked to answer a specific trial questionnaire with two randomly attributed versions, one for the experimental arm and one for the control arm. Briefly, the NutriNet-Santé cohort is an ongoing web-based prospective observational cohort study launched in France in May 2009, including adult volunteers recruited by multimedia campaigns, and aiming to study the relationships between diet and health, as well as the determinants of dietary behaviours (50).

Socio-demographic information were retrieved from the cohort questionnaire closest to the trial: sex, age, occupation, educational level, household income and area of residence (rural/urban). In both versions of the specific trial questionnaire, participants were first asked to self-estimate their nutrition knowledge level on a 4-point Likert scale (between ‘I am very knowledgeable about nutrition’ and ‘I do not know anything about nutrition’) and to self-evaluate the healthiness of their diet. Then, there were asked whether they had already heard about the Nutri-Score, through which source of information (television, radio, written and electronic press, governmental and institutional websites, social media, blogs, word of mouth, other), and whether what they had heard was rather positive or negative. They were then asked the same questions about the concept of “ultra-processed foods”. The complete trial questionnaire (two arms) is available online on the institutional website of the NutriNet-Santé cohort (51).

### Patient and public involvement

The research question underlying the study was driven by the motivation to improve citizens’ empowerment concerning their diets, by providing them with concrete tools to detect foods of higher nutritional quality and non-UPFs and allow them to make informed food choices. This corresponds to a strong demand from patients and consumers’ associations. Patients were not directly involved in the development of the protocol or in recruitment of participants. Dissemination of the research results will be done through the NutriNet-Santé cohort platform, including an abstract in French language, as well as through a press release and communication on social medias.

### Randomisation and blinding

Participants were randomly allocated to one of the two arms using a random drawing without replacement strategy. Given the nature of the intervention, participants could not be blinded of the intervention. However, they were only informed about the overall topic of the questionnaire, i.e., to study their objective understanding of two health dimensions of the diet: nutrient profile and food processing. Participants were not aware of the randomisation, neither of the presence of another arm or the explicit purpose of the trial.

### Experimental arm

The experimental arm consisted in the Nutri-Score 2.0 affixed on the front of all prepacked food products. The Nutri-Score V2.0 indicates 1) the overall nutrient profile of foods and beverages, using a 5-colour scale associated with letters—from dark green (A) to dark orange (E) for products (as calculated by the FSAm-NPS algorithm; details in the Supplementary Material, Appendix 1), and 2) in case the product is ultra-processed, a black banner surrounding the Nutri-Score, with the word “*ultra-transformé*”, meaning ultra-processed and no black banner otherwise (see Figure 1). Prior to the first webpage of trial, participants in the experimental arm had a short user notice, explaining how to interpret the Nutri-Score V2.0.

**Figure 1:**
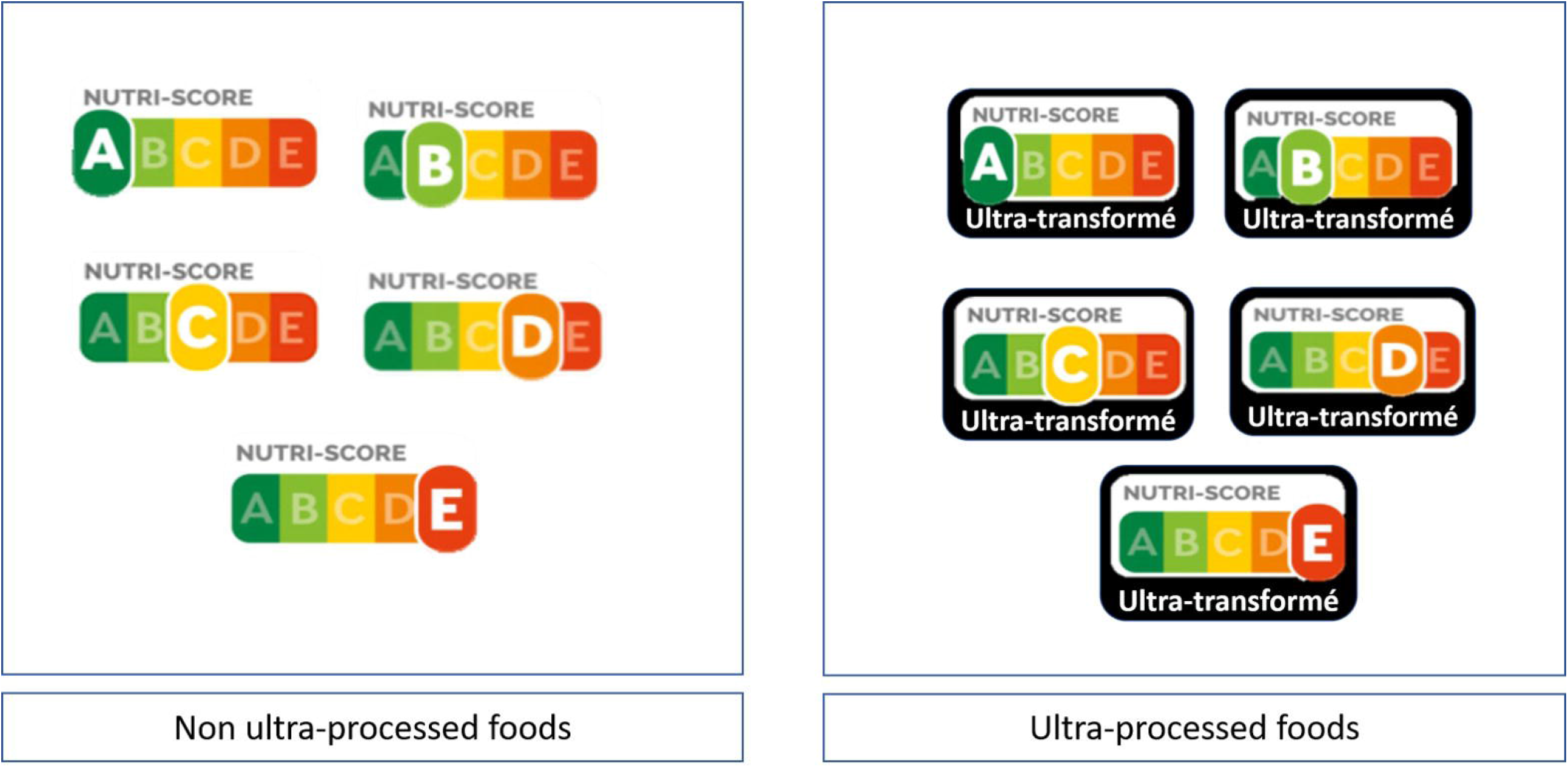
Conceptual design of the Nutri-Score 2.0

Participants received 3 sets of images of real food product packaging (brand blinded), categorized by food groups: 8 cookies, 7 breakfast cereals, and 7 ready-to-eat meals, with the Nutri-Score 2.0 displayed on the front-of-pack of each product. They had the possibility to check the back-of-pack nutrition facts and detailed list of ingredients (mandatory for food products on the EU market), by clicking on “I would like to turn the package around”, under each product. First, they were asked which product they would intend to purchase in each category, and which product they thought to be the “healthiest”. Then, participants were asked 1) to rank them according to their nutritional quality by identifying the first, the second and the third products with the best nutritional quality (in this order) and 2) to identify those that were ultra-processed. Last, a series of questions evaluated how participants of this arm perceived the Nutri-Score 2.0 and whether they found it helpful (details of each question in the Supplementary Material).

### Control arm

Participants received the same 3 sets of images of food packaging, but without front-of-pack nutritional label (in case the original Nutri-Score was displayed on the product, it was hidden by the investigators). As in the experimental arm, they had the possibility to check the back-of-pack nutrition facts and ingredient information. They were also asked, in each category, which product they would intend to purchase, and which product they thought to be the “healthiest”. Similarly, they were asked to rank them according to their nutritional quality and to identify those that were ultra-processed.

Both arms received the same definition of ultra-processed foods.

### Outcomes

#### Primary outcomes

In each food group (cookies, breakfast cereals, ready-to-eat meals), the primary outcome was the objective understanding of the two dimensions: nutritional quality (i.e., the nutrient profile) on the one hand and ultra-processing on the other. The number of correct answers was then counted. For nutritional quality, the answers of the participants were expected to match the order according to which Nutri-Score ranks the 3 products (highest nutritional quality, second highest, third highest). In case of ex-æquos (i.e., two products having the same Nutri-Score), both were considered correct. Therefore, the number of correct answers for the nutritional dimension could range between 0 (no correct answers) to 9 (3 best products*3 food categories, all correct answers).

For the processing dimension, participants were expected to identify all UPFs. For each product, the answer was considered correct if an UPF was identified as such by the participant (the gold standard being the presence of the black banner of the Nutri-Score 2.0, corresponding to the NOVA 4 “ultra-processed” definition (33)), and a non-UPF was identified as such. Therefore, the number of correct answers for the food processing dimension could range between 0 (no correct answers) and 22 (correct answer for all 22 products).

#### Secondary outcomes

Secondary outcomes were purchasing intentions (assessed with the question: “Which of these products would you purchase more frequently?”) and products perceived as the healthiest (assessed with the question “Which product seems the healthiest to you?”). The self-perceived ability of the participants to differentiate the nutrient profile of the products, as well as to detect UPFs was also assessed. The question for the experimental arm was: “Do you think that the Nutri-Score 2.0 helped you 1) differentiate the nutritional quality of these products?, 2) identify those that were ultra-processed?”; and for the control arm: “Did you feel able to 1) differentiate the nutritional quality of these products?, 2) identify those that were ultra-processed?”.

### Statistical analyses

The main objective of these questionnaires was to conduct the aforementioned randomised controlled trial; furthermore, a secondary aim was testing participants’ opinions and perceptions towards the suggested Nutri-Score 2.0 on a large sample size. Therefore, we planned a total sample size of 20,000 participants (10,000 per arm). Regarding the main objective, and considering an outcome prevalence of 10% in the control arm, this sample size would allow to detect as statistically significant an odds ratio estimate of 1.13 or 0.88 for primary outcomes, with an alpha (Type I) error of 5% and a power of 80%. To reach this final sample size while considering potential non-respondents, 30,000 NutriNet-Santé participants were randomly selected and were randomised in the two arms (n=15,000 for each). Socio-demographic characteristics of selected individuals who did and did not answer the questionnaires were compared using Chi-squared tests (eTable 1). Characteristics of the participants finally included in the trial were also compared between the two arms using Chi-squared tests. Questions on self-perceived knowledge on nutrition, self-perceived healthiness of the diet, as well as prior knowledge and sources of information on the original Nutri-Score and UPFs were compared between the two arms using Chi-squared tests.

For the primary outcomes, the numbers of correct answers were categorized into 3 classes: for the nutritional quality dimension: 1) 0-2 correct answers, 2) 3-6 correct answers, 3) 7-9 correct answers; for the ultra-processing dimension: 1) 0-12 correct answers, 2) 13-19 correct answers, 3) 20-22 correct answers. Sub-group analyses were additionally performed by considering the correct answers for each food category separately. The scores of correct answers for the 2 dimensions were described and compared between the two trial arms by Chi-squared tests. To measure the impact of the Nutri-Score 2.0 on the objective understanding of the 2 dimensions, multinomial logistic regression models (with the 3 categories of the score as outcome, the lowest category being the reference) were performed. To avoid potential residual confounding despite the randomization, these models were adjusted for age, sex, occupational status (active, inactive i.e., unemployed, student, retired), educational level (less than high school degree, <3 years after high school degree, ≥3 years after high school degree), household monthly income (less than 1100 euros, 1100-2330 euros, 2330-3780 euros, more than 3780 euros, I do not wish to answer) and area of residence (rural, urban). Interactions were tested in secondary analyses with educational level and self-reported knowledge in nutrition.

For secondary outcomes, purchasing intentions and products perceived as the healthiest were compared between the two arms with Chi-squared tests, same for the self-perceived ability of the participants to differentiate the nutritional quality of the products, as well as to detect UPFs.

Lastly, and in the experimental arm only, we described the answers of the participants regard their perception of the Nutri-Score 2.0, and whether they considered it helpful (these questions were specific to the experimental arm, by design).

We used the SQUIRE checklist when writing our report (52). All tests were two-sided, and a p value ≤0.05 was considered significant. Analyses were carried out with SAS software (V.9.4; SAS Institute).

## Results

### Characteristics of participants

Among the 30,000 NutriNet-Santé participants who received the trial questionnaire between April and June 2022 and who were randomised into one of the trial arms, 10,400 completed the questionnaire in the control arm, and 10,759 in the experimental arm (flowchart in appendix 2). Compared to participants who did not answer the questionnaire, included participants were more likely to be men, older, retired, and to have a lower educational level (eTable 1). Socio-demographic characteristics of the study population in the two arms were described in Table 1. The two arms did not differ according to any of the characteristics (all p-values >0.05). The sample consisted of 73% women, with a majority of participants (73%) older than 50 years old. Seventy percent of the sample had an educational level above high school, 68% were professionally inactive, 65% had a household monthly income higher than 2,330 euros and 76% lived in urban areas.

**Table 1:** Socio-demographic characteristics of included participants, according to arm of intervention, NutriNet-Santé, 2022, France, n = 21,159

|  | Control arm |  | Experimental arm |  | *p -value* |
| --- | --- | --- | --- | --- | --- |
|  | n | % | n | % |  |
|  | 10400 |  | 10759 |  |  |
| **Sex** |  |  |  |  |  |
| Women | 7621 | 73.3 | 7855 | 73.0 | 0.7 |
| Men | 2779 | 26.7 | 2904 | 27.0 |  |
| **Age category at date of questionnaire** |  |  |  |  |  |
| ]18-25 years] | 76 | 0.7 | 109 | 1.0 | 0.1 |
| ]25-50 years] | 2577 | 24.8 | 2752 | 25.6 |  |
| ]50-65 years] | 3507 | 33.7 | 3555 | 33.0 |  |
| >65 years | 4240 | 40.8 | 4343 | 40.4 |  |
| **Educational level** |  |  |  |  |  |
| < High school degree | 3146 | 30.3 | 3138 | 29.2 | 0.1 |
| <3 years after high school | 3255 | 31.3 | 3330 | 31.0 |  |
| ≥3 years after high school | 3999 | 38.5 | 4291 | 39.9 |  |
| **Professional situation** |  |  |  |  |  |
| Active | 4371 | 42.0 | 4678 | 43.5 | 0.2 |
| Inactive | 6029 | 58.0 | 6081 | 56.5 |  |
| **Household monthly income** |  |  |  |  |  |
| Less than 1110 euros/month | 267 | 2.6 | 259 | 2.4 | 0.7 |
| 1110 - 2330 euros/month | 1808 | 17.4 | 1922 | 17.9 |  |
| 2330 - 3780 euros/month | 2906 | 27.9 | 3017 | 28.0 |  |
| More than 3780 euros/month | 3865 | 37.2 | 4003 | 37.2 |  |
| Do not wish to answer | 1554 | 14.9 | 1558 | 14.5 |  |
| **Area of residence** |  |  |  |  |  |
| Unknown | 160 | 1.5 | 186 | 1.7 | 0.5 |
| Rural | 2286 | 22.0 | 2336 | 21.7 |  |
| Urban | 7954 | 76.5 | 8237 | 76.6 |  |

eTable 2 shows the two-arm comparisons regarding self-perceived knowledge on nutrition, self-perceived healthiness of the diet, as well as prior knowledge and sources of information on the original Nutri-Score and UPFs. Overall, no major differences were observed between the two arms. The large majority of the sample (more than 91%) considered themselves as having a healthy to very healthy diet. About 20% considered that they had none-to-low knowledge in nutrition. More than 95% of the participants in each arm had already heard about the original Nutri-Score prior to the study; television and the written or electronic press being the most frequently cited sources of information (58% and 50% respectively). More than 68% of the participants in each arm considered what they heard about Nutri-Score as rather positive to positive. On the other hand, more than 85% of the participants in each arm had already heard about UPFs prior to the study; television and the written or electronic press being also the most frequent sources of information (51% and 52% respectively). More than 82% of the participants in each arm declared that what they heard about UPFs was rather negative to very negative.

## Results for primary outcomes

Regarding the objective understanding of the nutritional quality of the products, 24.2% of the participants in the experimental arm obtained the highest possible scores (i.e. between 7 and 9 correct answers), vs. only 0.9% in the control arm. Specifically, these figures were 31.0% vs. 5.5% for cookies, 26.0% vs. 1.0% for breakfast cereals, and 23.5 vs. 1.3% for ready-to-eat meals (Table 2, all P<0.0001).

**Table 2:**
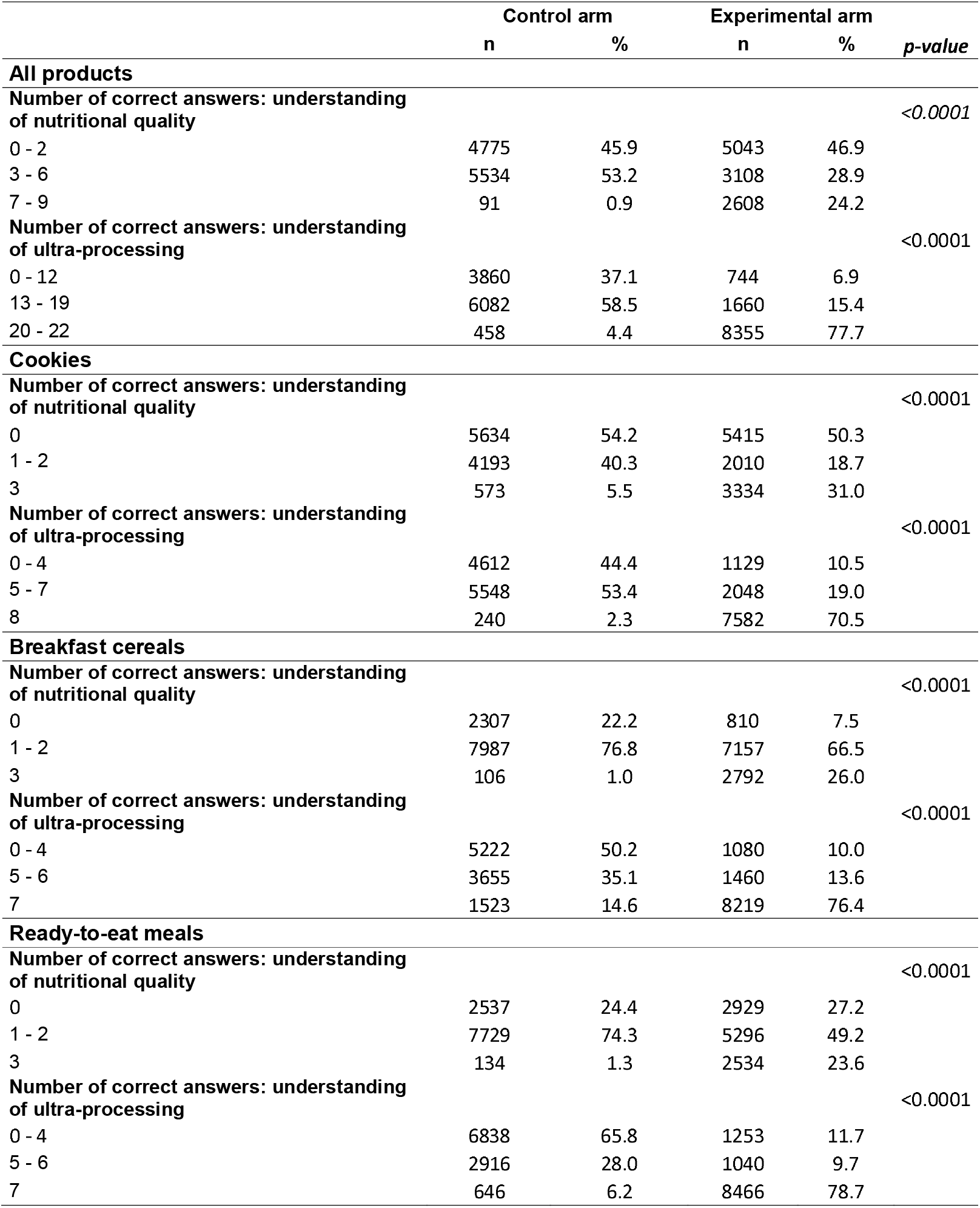
Number of correct answers according to arm of intervention, NutriNet-Santé, 2022, France n = 21,159

|  | Control arm |  | Experimental arm |  |  |
| --- | --- | --- | --- | --- | --- |
|  | n | % | n | % | p-value |
| All products |  |  |  |  |  |
| Number of correct answers: understanding of nutritional quality |  |  |  |  | <0.0001 |
| 0 - 2 | 4775 | 45.9 | 5043 | 46.9 |  |
| 3 - 6 | 5534 | 53.2 | 3108 | 28.9 |  |
| 7 - 9 | 91 | 0.9 | 2608 | 24.2 |  |
| Number of correct answers: understanding of ultra-processing |  |  |  |  | <0.0001 |
| 0 - 12 | 3860 | 37.1 | 744 | 6.9 |  |
| 13 - 19 | 6082 | 58.5 | 1660 | 15.4 |  |
| 20 - 22 | 458 | 4.4 | 8355 | 77.7 |  |
| Cookies |  |  |  |  |  |
| Number of correct answers: understanding of nutritional quality |  |  |  |  | <0.0001 |
| 0 | 5634 | 54.2 | 5415 | 50.3 |  |
| 1 - 2 | 4193 | 40.3 | 2010 | 18.7 |  |
| 3 | 573 | 5.5 | 3334 | 31.0 |  |
| Number of correct answers: understanding of ultra-processing |  |  |  |  | <0.0001 |
| 0 - 4 | 4612 | 44.4 | 1129 | 10.5 |  |
| 5 - 7 | 5548 | 53.4 | 2048 | 19.0 |  |
| 8 | 240 | 2.3 | 7582 | 70.5 |  |
| Breakfast cereals |  |  |  |  |  |
| Number of correct answers: understanding of nutritional quality |  |  |  |  | <0.0001 |
| 0 | 2307 | 22.2 | 810 | 7.5 |  |
| 1 - 2 | 7987 | 76.8 | 7157 | 66.5 |  |
| 3 | 106 | 1.0 | 2792 | 26.0 |  |
| Number of correct answers: understanding of ultra-processing |  |  |  |  | <0.0001 |
| 0 - 4 | 5222 | 50.2 | 1080 | 10.0 |  |
| 5 - 6 | 3655 | 35.1 | 1460 | 13.6 |  |
| 7 | 1523 | 14.6 | 8219 | 76.4 |  |
| Ready-to-eat meals |  |  |  |  |  |
| Number of correct answers: understanding of nutritional quality |  |  |  |  | <0.0001 |
| 0 | 2537 | 24.4 | 2929 | 27.2 |  |
| 1 - 2 | 7729 | 74.3 | 5296 | 49.2 |  |
| 3 | 134 | 1.3 | 2534 | 23.6 |  |
| Number of correct answers: understanding of ultra-processing |  |  |  |  | <0.0001 |
| 0 - 4 | 6838 | 65.8 | 1253 | 11.7 |  |
| 5 - 6 | 2916 | 28.0 | 1040 | 9.7 |  |
| 7 | 646 | 6.2 | 8466 | 78.7 |  |

As regards the objective understanding of ultra-processing, 77.7% of the participants in the experimental arm obtained the highest possible scores (i.e., between 20 and 22 correct answers), vs. only 4.4% in the control arm. These figures were 70.5% vs. 2.3% for cookies, 76.4% vs. 14.6% for breakfast cereals, and 78.7 vs. 6.2% for ready-to-eat meals (Table 2, all P<0.0001).

In logistic regression models (Table 3), the Nutri-Score 2.0 led to higher odds of getting the highest scores, for both studied food dimensions: OR = 29.0 (23.4 – 35.9), p<0.001 for nutritional quality (the highest scores being 7-9 correct answers), and OR = 174.3 (151.4 – 200.5), p<0.001 for ultra-processing (the highest scores being 20-22 correct answers). The results were similar across the 3 food categories (cookies, breakfast cereals, and ready-to-eat meals).

**Table 3:** Impact of the Nutri-Score 2.0 on primary outcomes (i.e., objective understanding of nutritional quality and food ultra-processing), NutriNet-Santé, 2022, France, n = 21,159

|  | OR* (95% CI) [experimental arm vs. control arm] |  |  | P-trend |
| --- | --- | --- | --- | --- |
| **All products** |  |  |  |  |
| **Number of correct answers** | 0 - 2 | 3 - 6 | 7 - 9 |  |
| Understanding of nutritional quality | 1 | 0.54 (0.51 - 0.57) | 29.0 (23.4 - 35.9) | <0.0001 |
| **Number of correct answers** | 0 - 12 | 13 - 19 | 20 - 22 |  |
| Understanding of ultra-processing | 1 | 1.94 (1.75 - 2.14) | 174.3 (151.4 - 200.5) | <0.0001 |
| **Cookies** |  |  |  |  |
| **Number of correct answers** | 0 | 1 - 2 | 3 |  |
| Understanding of nutritional quality | 1 | 0.50 (0.47 - 0.53) | 6.46 (5.86 - 7.13) | <0.0001 |
| **Number of correct answers** | 0 - 4 | 5 - 7 | 8 |  |
| Understanding of ultra-processing | 1 | 1.89 (1.73 - 2.06) | 191.4 (164.2 - 223.1) | <0.0001 |
| **Breakfast cereals** |  |  |  |  |
| **Number of correct answers** | 0 | 1 - 2 | 3 |  |
| Understanding of nutritional quality | 1 | 2.75 (2.51 - 3.00) | 86.8 (70.2 - 107.3) | <0.0001 |
| **Number of correct answers** | 0 - 4 | 5 - 6 | 7 |  |
| Understanding of ultra-processing | 1 | 2.37 (2.16 - 2.60) | 35.6 (32.4 - 39.2) | <0.0001 |
| **Ready-to-eat meals** |  |  |  |  |
| **Number of correct answers** | 0 | 1 - 2 | 3 |  |
| Understanding of nutritional quality | 1 | 0.59 (0.56 - 0.63) | 17.3 (14.4 - 20.8) | <0.0001 |
| **Number of correct answers** | 0 - 4 | 5 - 6 | 7 |  |
| Understanding of ultra-processing | 1 | 2.38 (2.16 - 2.62) | 100.0 (89.5 - 111.7) | <0.0001 |

**OR\* = Odds Ratio**
ORs derived from multinomial logistic regression models to predict the number of correct answers according to the experimentation arm, adjusted for age, sex, educational level, household monthly income, professional situation and area of residence

Even though interactions tests were significant with educational level and self-reported knowledge in nutrition, trends were similar in stratified analyses. Of note, the effect magnitude of the label was approximately twice as stronger in participants reporting being less knowledgeable about nutrition, compared to those reporting being well knowledgeable (eTable 3).

### Results for secondary outcomes

As regards secondary outcomes (eTable 4), participants in the experimental arm were more likely than those in the control arm to prefer as purchasing intentions foods with the both higher nutritional quality and non-ultra-processed products (all p-values <0.001). In case of a discordance between the nutritional quality and food ultra-processing (i.e., if the product with the best nutritional quality (Nutri-Score A or B) is ultra-processed, with no non-ultra-processed alternative with the same Nutri-Score, as it was the case for cookies and ready-to-eat meals), participants in the Nutri-Score 2.0 arm generally tended to privilege the non-ultra-processed product having the best available Nutri-Score rather than an UPF having a better Nutri-Score (eTable 4).

The impact of Nutri-Score 2.0 on the products perceived as the healthiest followed the same trends as purchasing intentions (eTable 4). Compared with participants in the control arm, those in the experimental arm reported being more frequently able to differentiate the nutritional quality of cookies (77.5% vs. 58.9), breakfast cereals (84.7% vs. 50.3%), and ready-to-eat meals (80.4% vs. 38.7%) (all p-values <0.001). They also reported being more frequently able to identify ultra-processed foods among cookies (91.9% vs. 52.0%), breakfast cereals (91.9% vs. 50.0%), and ready-to-eat meals (90.2% vs. 42.8%) (all p-values <0.001) (eTable 4).

Lastly, as shown in eTable 5, participants in the experimental arm had a positive perception of the Nutri-Score 2.0: more than three quarters of the participants in this arm found that the Nutri-Score 2.0 was credible, trustworthy and easy to understand. They also reported that Nutri-Score 2.0 would help them with their purchases (84.1% declared that they rather agreed or totally agreed), would give them useful information for food purchases (85.5%), would help them to differentiate the nutritional quality of food products (82.5%) and to identify UPFs (93.9%). Finally, 88.0% of the participants would like to see the Nutri-Score 2.0 on the front-of-pack of food products.

## Discussion

This randomised experimental study showed that a graphically updated version of the Nutri-Score including an additional graphical banner mentioning “Ultra-processed” for UPFs (Nutri-Score 2.0) increased significantly the objective understanding of both the nutrient profile dimension of food products, and the ultra-processing dimension, compared with no front-of-pack label. The latter corresponds, in accordance with the European regulations in vigour, to the current official situation in Europe since no front-of-pack nutrition label is mandatory at the moment. The Nutri-Score 2.0 also had a positive impact on purchasing intentions and on the products perceived as the healthiest, guiding consumers towards a better nutrient profile and non-ultra-processed products. Participants in the Nutri-Score 2.0 arm had in majority a positive perception of the label, and found it useful and trustworthy.

To our knowledge, no study previously investigated the impact of a front-of-pack label combining nutrient profile and food ultra-processing on the objective understanding of these dimensions, or on purchasing intensions and the products perceived as the healthiest.

However, these results were in line with previously published studies (2,11,53–56) showing that interpretive front-of-pack labels, including the Nutri-Score, were associated with a large improvement in the ability of participants to correctly rank foods according to their nutritional quality, even though a direct comparison of effect sizes would not be straightforward. This positive effect may be partly linked, in the case of the Nutri-Score, to its color-coding scheme, using the green-dark orange polychromatic scale, which might reduce the time needed by participants to identify and integrate the nutritional information (57). Our results on the impact on purchasing intentions were also consistent with previously published studies showing that interpretive front-of-pack labels have a potential positive effect on the nutritional quality of consumers’ choice in studies based on questionnaires, randomised trials and experimental studies in virtual supermarkets (5,11,58–61), and studies observing higher performance of summary labels to improve food choices (60,62–64).

All these studies investigated the effects of the labels on the objective understanding / purchasing intentions related to the product’s nutrient profile, i.e., in line with the rationale behind these labels. In the present study, the Nutri-Score 2.0 had an impact on the objective understanding of both the nutrient profile dimension and the food ultra-processing dimension, showing that these two complementary dimensions could be independently perceived and understood by the participants. Interestingly, we observed higher odds ratios for the ultra-processing dimension compared with the nutrient profile dimension, even though this comparison is not straightforward. The most obvious explanation could be linked to the different complexity levels of both dimensions; the nutrient profile being a multi-level and relative (used to compare similar products) dimension, while ultra-processing is a binary absolute dimension (i.e., no need for a comparison to tell that a product is ultra-processed or not). In addition, the task requested from participants was more complicated for the nutrient profile (i.e., ranking) compared with that of ultra-processing (i.e., a binary outcome). Even though trends were mostly similar across the 3 studied food categories, the effect magnitude was different: as regards the nutritional quality, the Nutri-Score 2.0 had the strongest effect for breakfast cereals, followed by ready-to-eat meals then cookies; while for ultra-processing, the effect was the strongest for cookies, followed by ready-to-eat meals then breakfast cereals. As regards the impact of the Nutri-Score 2.0 on the objective understanding of the nutrient profile dimension, we observed J-shaped associations specifically in cookies and ready-to-eat meals. This could be explained by the selected products within these categories: indeed, these series of products presented a “discordance” between the nutritional quality and food ultra-processing (i.e., the product with the best nutrient profile (Nutri-Score A or B) was ultra-processed, without any non-ultra-processed alternative with the same Nutri-Score). This design was intentional, in order to study how consumers would spontaneously arbitrate between these two complementary dimensions, even in a context where science is unable so far to conclude on which of the two dimensions has a higher impact on long term human health. Participants in the Nutri-Score 2.0 arm seemed to occasionally overestimate the nutrient profile of some non-ultra-processed products, as they tended to rank non-ultra-processed products with a lower nutritional value ahead of ultra-processed products with a higher nutritional value. This was also in line with our analyses on purchasing intentions and the products perceived as the healthiest, showing that in the Nutri-Score 2.0 arm, when participants encountered this aforementioned “discordant” situation, they were more inclined to select the non-ultra-processed products with a worse nutrient profile as a purchasing intention and to subjectively perceive it as a healthier product. The cognitive arbitration of the participants towards the food ultra-processing dimension might be linked to the extensive recent studies published on this topic, to the resulting mediatic hype that UPFs have received, and to the recent inclusion of limits for consumption of UPFs in French dietary guidelines (65), while well-scientifically established nutritional concepts (e.g. harmful effects of excessive sugar, salt, low fiber intake, etc.) may have received less attention from the press and public lately in the French context. However, we cannot exclude that the graphical difference between the two dimensions (especially with the black colour attributed to UPFs in this prototype) could have influenced the participants into perceiving the ultra-processing dimension as a more important one. Nonetheless, scientific evidence illustrates the independent health impacts of both nutrient profile and food processing and the support the idea that consumers should be transparently informed on these two complementary dimensions (29,66).

This study presents several strengths: the randomised controlled trial design resulted in socio-demographically comparable groups and limitation of confounding bias. Second, the online architecture of the NutriNet-Santé cohort and the emailing system to include participants allowed achieving a robust statistical power. Finally, participants had also access to the mandatory back-of-pack nutrition facts and ingredient information, increasing the external validity of the study (same level of information as online supermarkets). However, limitations should be acknowledged. First, participants were recruited among a volunteer-based cohort on nutrition and health. Therefore, participants may have been more interested in nutrition than the general French population (67) (more than 91% considered their diet as healthy to very healthy). Thus, the gain brought by a simplified nutritional logo may have been minimized in this study population relatively interested and accustomed to seeking information on the packaging of the products they consume. Furthermore, the number of products was relatively limited, and participants were sometimes not interested in any of the products (between 27 and 52% of the participants reported not willing to buy any of the suggested products, within each category). This might have limited the interest of the participants in the study, which could potentially result in classification bias. Another related limitation is the observed opt-in phenomenon in the experimental arm (less participants declared willing to purchase “none of the products”, in the experimental arm). It would have been possible that this label has encouraged participants in the experimental arm to purchase products they would have never intended to purchase without the label, through a potential positivity bias (68); yet, this assumption could not be verified using our design, and would need a cross-over trial. This potential opt-in phenomenon might be a limitation in case of unfavourable food groups, such as cookies (e.g. 6% less participants in the experimental arm reporting intending to purchase ‘none of the suggested cookies’), especially considering the fact that the product more frequently intended to be purchased by the participants in the experimental arm (Nutri-Score C, not ultra-processed) did not correspond to the one with the best nutrient profile (Nutri-Score A, ultra-processed). However, this worse nutrient profile might be compensated by a decrease of UPF intakes, to an extent that is not fully understood yet. The implications of these purchasing intentions on the overall quality of the diet (according to both the nutrient profile and ultra-processing) need further investigations with complementary study designs (e.g. virtual or experimental supermarkets). Next, the Nutri-Score 2.0 was accompanied by a pedagogic notice prior to the trial webpage, explaining how this label should be used. This is not systematically the case in reality, as communication campaigns about front-of-pack labels might be insufficient or not at the reach of the whole population. This could have overestimated our associations, even though the majority of participants in the two arms declared being familiar with both the Nutri-Score and the concept of ultra-processing. Moreover, we hypothesise that the implementation of such labels should always be accompanied by massive educational communication campaigns. In addition, other labels such as the organic label were not hidden from the packages and were not randomised. This could have misled the participants into selecting the organic products as the healthier ones, but this situation is the same in real-life. Lastly, only one graphical format combining the two dimensions has been tested. Further studies comparing different graphical shapes and colours for the “UPF” indication would be interesting to perform, as well as crossover studies and discrete choice experiments exploring the effect of the opt-in phenomenon resulting from this label among different food groups.

In conclusion, this randomised controlled trial demonstrates the interest of a front-of-pack logo combining the NutriScore (informing on the nutrient profile dimension) with an additional graphic mention indicating whether the food is ultra-processed or not, compared to a no-label situation. This “NutriScore 2.0” strongly improved the ability of the participants to detect food with a better nutrient profile and to identify UPFs compared to the current official situation in Europe, i.e. no mandatory front-of-pack nutrition or UPF label. This label had also an impact on purchasing intentions and the products perceived as the healthiest, towards a better nutritional quality and a reduction of UPFs. Studies investigating the impact of this label in different subgroups of populations should be performed, as well as studies on the effect of the Nutri-Score 2.0 on purchasing intentions of other food categories. Adding information regarding the food processing dimension to interpretive front-of-pack nutritional labels might be of public health interest for consumers, as our results show that they could be able to independently identify and understand these two correlated, but distinct and complementary dimensions.

## Data Availability

Data described in the manuscript, code book, and analytic code will be made available upon request pending application and approval. Researchers from public institutions can submit a collaboration request including information on the institution and a brief description of the project to. All requests will be reviewed by the steering committee of the NutriNet-Sante study. If the collaboration is accepted, a data access agreement will be necessary and appropriate authorizations from the competent administrative authorities may be needed. In accordance with existing regulations, no personal data will be accessible.

## Data availability

Data described in the manuscript, code book, and analytic code will be made available upon request pending application and approval. Researchers from public institutions can submit a collaboration request including information on the institution and a brief description of the project to. All requests will be reviewed by the steering committee of the NutriNet-Santé study. If the collaboration is accepted, a data access agreement will be necessary and appropriate authorizations from the competent administrative authorities may be needed. In accordance with existing regulations, no personal data will be accessible.

## Ethics approval

The study was approved by the Institutional Review Board of Inserm (IRB Inserm n°IRB0000388 FWA00005831) and the National Commission for Data Protection and Liberties (CNIL n°909216), and registered at: https://clinicaltrials.gov/show/NCT05610930. Electronic consent was obtained from all participants. The NutriNet-Santé study is conducted according to the Declaration of Helsinki guidelines, approved by the IRB of Inserm (n°0000388FWA00005831) and the National Commission for Data Protection and Liberties (n°908450/n°909216), and registered at: https://clinicaltrials.gov/ct2/show/NCT03335644.

## Acknowledgements

We thank Thi Hong Van Duong, Régis Gatibelza, Jagatjit Mohinder, Rizvane Mougamadou, and Aladi Timera (computer scientists); Julien Allegre, Nathalie Arnault, Nicolas Dechamp, (data-manager/statisticians); Paola Yvroud (health event validator); Maria Gomes and Mirette Foham (Nutrinaute support) for their technical contribution to the NutriNet-Santé study. We warmly thank all the volunteers of the NutriNet-Santé cohort.

## Contributorship statement and guarantor

The authors’ contributions were as follows – BSr, SH, PG, EKG, CJ and MT designed the experiment. NDP and YE coordinated data collection. BSr and MT designed the analysis plan. FSE and LB conducted statistical analyses. BSr supervised statistical analyses. BSr wrote the first draft approved by all authors, and revised the manuscript. MT supervised the writing. CJ and MT contributed equally, and share last authorship. All authors: contributed to the data interpretation, revised each draft for important intellectual content and approved the final manuscript. BSr and MT had primary responsibility for the final content, they are the guarantors. The corresponding author (BSr) attests that all listed authors meet authorship criteria and that no others meeting the criteria have been omitted.

## Transparency statement

The guarantors affirm that the manuscript is an honest, accurate, and transparent account of the study being reported; that no important aspects of the study have been omitted; and that any discrepancies from the study as planned have been explained.

## Supplementary material

**Appendix 1:** FSAm-NPS algorithm calculation

**eTable 1:** Comparison of socio-demographic participants between participants who answered the questionnaire (included) and those who were selected but did not answer

**Appendix 2:** Flowchart for sample selection, NutriNet-Santé, France, 2022, n = 21,159

**eTable 2:** Participants’ knowledge about nutrition, Nutri-Score and food ultra-processing, NutriNet-Santé, 2022, France, n = 21,159

**eTable 3:** Impact of the Nutri-Score 2.0 on primary outcomes (i.e., objective understanding of nutritional quality and food ultra-processing) in stratified analyses, NutriNet-Santé, 2022, France, n = 21,159

**eTable 4:** Secondary outcomes: comparison of the 2 arms in terms of purchasing intensions and the products perceived as the healthiest, NutriNet-Santé, 2022, France, n = 21,159

**eTable 5:** Perception of the Nutri-Score 2.0 by the experimental arm participants, NutriNet-Santé, 2022, France, n = 10,759

