## Supplemental material for "Effect of a new graphically modified Nutri-Score on the objective understanding of foods’ nutrient profile and ultra-processing – a randomised controlled trial"

**Supplementary material**

**Appendix 1:** FSAm-NPS algorithm calculation

Points are allocated according to the nutrient content for 100g of foods or beverages.

Points are allocated for ‘Negative’ nutrients (A points) and can be balanced according to ‘Positive’ nutrients (C points).

***A points***

Total A points = (points for energy) + (points for saturated fat) + (points for total sugar) + (points for sodium)

| ***Points*** | **Energy (kJ)** | **Saturated Fat (g)** | **Total Sugars (g)** | **Sodium (mg)** |
| --- | --- | --- | --- | --- |
| 0 | ≤ 335 | ≤ 1 | ≤ 4.5 | ≤ 90 |
| 1 | > 335 | > 1 | > 4.5 | > 90 |
| 2 | > 670 | > 2 | > 9 | > 180 |
| 3 | > 1005 | > 3 | > 13.5 | > 270 |
| 4 | > 1340 | > 4 | > 18 | > 360 |
| 5 | > 1675 | > 5 | > 22.5 | > 450 |
| 6 | > 2010 | > 6 | > 27 | > 540 |
| 7 | > 2345 | > 7 | > 31 | > 630 |
| 8 | > 2680 | > 8 | > 36 | > 720 |
| 9 | > 3015 | > 9 | > 40 | > 810 |
| 10 | > 3350 | > 10 | > 45 | > 900 |

***C points***

Total C points = (points for fruits/vegetables/legumes/nuts) + (points for fibres) + (points for proteins)

| ***Points*** | **Fruits/vegetables/legumes/nuts** **(%)** | **Fibre (g) *** | **Protein (g)** |
| --- | --- | --- | --- |
| 0 | ≤ 40 | ≤ 0.7 | ≤ 1.6 |
| 1 | > 40 | > 0.7 | > 1.6 |
| 2 | > 60 | > 1.4 | > 3.2 |
| 3 | - | > 2.1 | > 4.8 |
| 4 | - | > 2.8 | > 6.4 |
| 5 | > 80 | > 3.5 | > 8.0 |

* FSAm-NPS score allocates different thresholds for fibres, depending on the measurement method used. We used NSP cut-offs to compute fibres score.

For 100g of a given food, the percentage of fruits/vegetables/legumes/nuts is obtained by summing up the amount (in grams) of all fruits, legumes and vegetables (including oleaginous fruits, dried fruits and olives) contained in this food.

***Overall score computation***

- If Total A points <11, then FSAm-NPS score =Total A points – Total C points
- If Total A points ≥11,
  - If points for fruits/vegetables/legumes/nuts =5, then FSAm-NPS score =Total A points – Total C points
  - Else if points for fruits/vegetables/legumes/nuts <5, then FSAm-NPS score = Total A points – (points for fibre + points for fruits/vegetables/legumes/nuts).

Exceptions were made for cheese, added fat, and drinks to better rank them according to their nutrient profile, consistently with nutritional recommendations:

***Score computation for cheese***

For cheese, the score takes in account the protein content, whether the A score reaches 11 or not, i.e.: FSAm-NPS score =Total A points – Total C points

***Score computation for added fat***

For added fat, the grid for point attribution is based on the percentage of saturated fat among total lipids (instead of saturated fat (g)) and has a six-point homogenous ascending step, as shown thereafter:

| ***Points*** | **Saturated Fat/Lipids (%)** |
| --- | --- |
| 0 | < 10 |
| 1 | < 16 |
| 2 | < 22 |
| 3 | < 28 |
| 4 | < 34 |
| 5 | < 40 |
| 6 | < 46 |
| 7 | < 52 |
| 8 | < 58 |
| 9 | < 64 |
| 10 | ≥ 64 |

Points attribution for the other nutrients follows the grid displayed in “A points” and “C points” above.

***Score computation for drinks***

For drinks, the grids for point attribution regarding energy, sugars and fruits/vegetables/ legumes/nuts (%) were modified.

| ***Points*** | **Energy (kJ)** | **Sugars (g)** | **Fruits/vegetables/legumes/nuts (%)** |
| --- | --- | --- | --- |
| 0 | ≤ 0 | ≤ 0 | < 40 |
| 1 | ≤ 30 | ≤ 1.5 |  |
| 2 | ≤ 60 | ≤ 3 | > 40 |
| 3 | ≤ 90 | ≤ 4.5 |  |
| 4 | ≤ 120 | ≤ 6 | > 60 |
| 5 | ≤ 150 | ≤ 7.5 |  |
| 6 | ≤ 180 | ≤ 9 |  |
| 7 | ≤ 210 | ≤ 10.5 |  |
| 8 | ≤ 240 | ≤ 12 |  |
| 9 | ≤ 270 | ≤ 13.5 |  |
| 10 | > 270 | > 13.5 | > 80 |

Points attribution for the other nutrients follows the grid displayed in “A points” and “C points” above.

Given the modification of the grid for fruit and vegetables for beverages, the threshold in the final computation to take into account protein content is set at 10 points:

- If Total A points <11, then FSAm-NPS score =Total A points – Total C points
- If Total A points ≥11,
  - If points for fruits/vegetables/legumes/nuts =10, then FSAm-NPS score =Total A points – Total C points
  - Else if points for fruits/vegetables/legumes/nuts <10, then FSAm-NPS score = Total A points – (points for fibre + points for fruits/vegetables/legumes/nuts).

Milk and vegetable milk are not concerned by this exception. Their scores are computed using the overall score computation system.

***FSAm-NPS score and Attribution of Nutri-Score colours***

| **Foods (points)** | **Beverages (points)** | **Colour** | |
| --- | --- | --- | --- |
| Min to -1 | Water | Dark green | *Highest nutritional quality* |
| 0 to 2 | Min to 1 | Light green |  |
| 3 to 10 | 2 to 5 | Yellow |  |
| 11 to 18 | 6 to 9 | Light orange |  |
| 19 to max | 10 to max | Dark orange | *Lowest nutritional quality* |

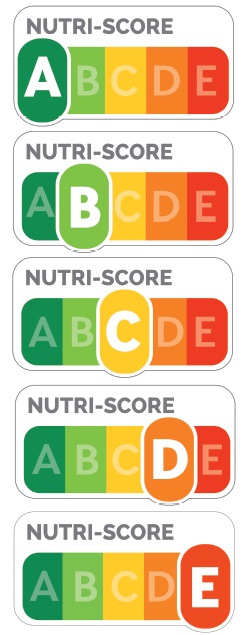

Santé Publique France 2017, Nutri-Score Logo

**eTable 1:** Comparison of socio-demographic participants between participants who answered the questionnaire (included) and those who were selected but did not answer, NutriNet-Santé, 2022, n = 30,000

|  | **Included** | | **Non-included** | |  |
| --- | --- | --- | --- | --- | --- |
|  | **n** | **%** | **n** | **%** | ***p -value*** |
|  | 21159 |  | 7888 |  |  |
| **Sex** |  |  |  |  |  |
| Women | 15476 | 73.1 | 6191 | 78.5 |  |
| Men | 5683 | 26.9 | 1697 | 21.5 | <0.0001 |
| **Age category at date of questionnaire** |  |  |  |  |  |
| ]18-25 years] | 185 | 0.9 | 134 | 1.7 |  |
| ]25-50 years] | 5329 | 25.2 | 2787 | 35.3 |  |
| ]50-65 years] | 7062 | 33.4 | 2568 | 32.6 |  |
| >65 years | 8583 | 40.6 | 2399 | 30.4 | <0.0001 |
| **Educational level** |  |  |  |  |  |
| < High school degree | 6284 | 29.7 | 1981 | 25.1 |  |
| <3 years after high school | 6585 | 31.1 | 2538 | 32.2 |  |
| ≥3 years after high school | 8290 | 39.2 | 3369 | 42.7 | <0.0001 |
| **Professional situation** |  |  |  |  |  |
| Unemployed | 1328 | 6.3 | 585 | 7.4 |  |
| Student | 155 | 0.7 | 121 | 1.5 |  |
| Retired | 10627 | 50.2 | 2940 | 37.3 |  |
| Employees, agriculture professions, workers | 2244 | 10.6 | 1085 | 13.8 |  |
| Intermediate and superior professions | 6805 | 32.2 | 3157 | 40.0 | <0.0001 |
| **Household monthly income** |  |  |  |  |  |
| Less than 1110 euros/month | 526 | 2.5 | 191 | 2.4 |  |
| 1110 - 2330 euros/month | 3730 | 17.6 | 1258 | 16.0 |  |
| 2330 - 3780 euros/month | 5923 | 28.0 | 2123 | 26.9 |  |
| More than 3780 euros/month | 7868 | 37.2 | 3012 | 38.2 |  |
| Do not wish to answer | 3112 | 14.7 | 1304 | 16.5 | <0.0001 |
| **Area of residence** |  |  |  |  |  |
| Unknown | 4639 | 21.9 | 1745 | 22.1 |  |
| Rural | 16175 | 76.5 | 6003 | 76.1 |  |
| Urban | 345 | 1.6 | 140 | 1.8 | 0.6 |

**Appendix 2:** Flowchart for sample selection, NutriNet-Santé, 2022, n = 21,159

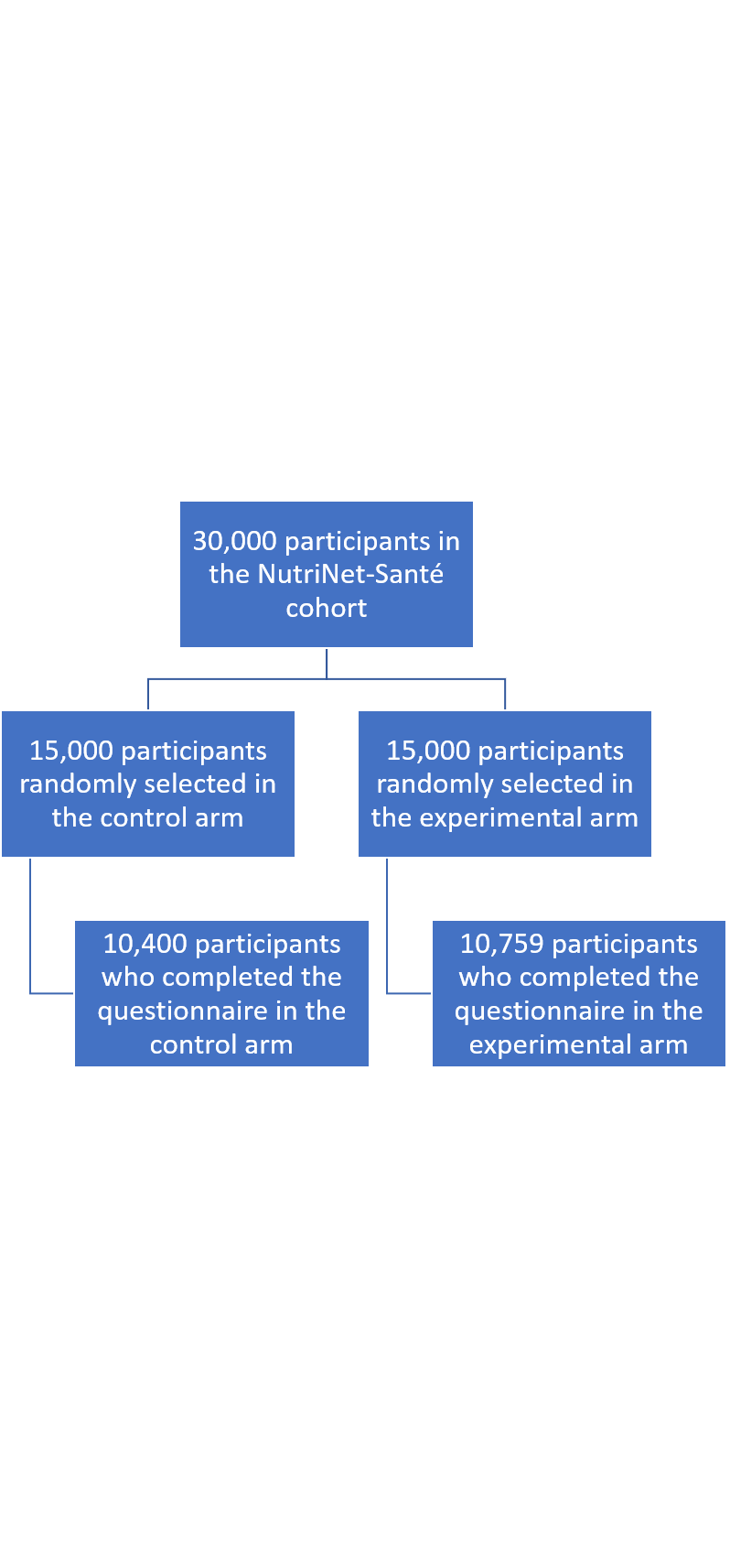

**eTable 2:** Participants’ knowledge about nutrition, Nutri-Score and food ultra-processing, NutriNet-Santé, 2022, n = 21,159

|  | **Control arm** | | **Experimental arm** | |  |
| --- | --- | --- | --- | --- | --- |
|  | **n** | **%** | **n** | **%** | ***p-value*** |
|  | 10400 |  | 10759 |  |  |
| **How do you consider your diet?** |  |  |  |  |  |
| Very unhealthy | 56 | 0.5 | 76 | 0.7 |  |
| Unhealthy | 612 | 5.9 | 668 | 6.2 |  |
| Healthy | 7980 | 76.7 | 8148 | 75.7 |  |
| Very healthy | 1673 | 16.1 | 1767 | 16.4 |  |
| I do not know | 79 | 0.8 | 100 | 0.9 | 0.2 |
| **How do you estimate your knowledge in nutrition?** |  |  |  |  |  |
| I do not know anything about nutrition | 107 | 1.0 | 90 | 0.8 |  |
| I know few things about nutrition | 2028 | 19.5 | 1988 | 18.5 |  |
| I am rather knowledgeable about nutrition | 6608 | 63.5 | 6932 | 64.4 |  |
| I am very knowledgeable about nutrition | 1484 | 14.3 | 1562 | 14.5 |  |
| I do not know | 173 | 1.7 | 187 | 1.7 | 0.2 |
| **Prior to this questionnaire, had you heard about the Nutri-Score in the press or on the Internet?** |  |  |  |  |  |
| No | 479 | 4.6 | 458 | 4.3 |  |
| Yes | 9921 | 95.4 | 10301 | 95.7 | 0.2 |
| **From where had you heard about it?** |  |  |  |  |  |
| On television | 6047 | 58.1 | 6321 | 58.8 | 0.4 |
| On the radio | 2955 | 28.4 | 3033 | 28.2 | 0.7 |
| In written an electronic press | 5265 | 50.6 | 5451 | 50.7 | 1.0 |
| On an institutional or governmental website | 3398 | 32.7 | 3502 | 32.6 | 0.9 |
| On social media | 1073 | 10.3 | 1134 | 10.5 | 0.6 |
| On a blog | 147 | 1.4 | 143 | 1.3 | 0.6 |
| Word of mouth | 1280 | 12.3 | 1473 | 13.7 | 0.003 |
| Other | 1505 | 14.5 | 1593 | 14.8 | 0.5 |
| **What you had heard about the Nutri-Score was:** |  |  |  |  |  |
| Very negative | 38 | 0.4 | 33 | 0.3 |  |
| Rather negative | 727 | 7.0 | 788 | 7.3 |  |
| Neutral | 2067 | 19.9 | 2125 | 19.8 |  |
| Rather positive | 6193 | 59.6 | 6479 | 60.2 |  |
| Very positive | 896 | 8.6 | 876 | 8.1 |  |
| I do not know | 479 | 4.6 | 458 | 4.3 | 0.5 |
| **Prior to this questionnaire, had you heard about the concept of ultra-processed foods?** |  |  |  |  |  |
| No | 1502 | 14.4 | 1568 | 14.6 |  |
| Yes | 8898 | 85.6 | 9191 | 85.4 | 0.8 |
| **From where had you heard about it?** |  |  |  |  |  |
| On television | 5373 | 51.7 | 5458 | 50.7 | 0.2 |
| On the radio | 3289 | 31.6 | 3290 | 30.6 | 0.1 |
| In written an electronic press | 5424 | 52.2 | 5592 | 52.0 | 0.8 |
| On an institutional or governmental website | 2141 | 20.6 | 2204 | 20.5 | 0.9 |
| On social media | 1059 | 10.2 | 1203 | 11.2 | 0.02 |
| On a blog | 241 | 2.3 | 274 | 2.6 | 0.3 |
| Word of mouth | 1327 | 12.8 | 1507 | 14.0 | 0.008 |
| Other | 1254 | 12.1 | 1334 | 12.4 | 0.5 |
| **What you had heard about ultra-processed foods was:** |  |  |  |  |  |
| Very negative | 6456 | 62.1 | 6641 | 61.7 |  |
| Rather negative | 2107 | 20.3 | 2207 | 20.5 |  |
| Neutral | 144 | 1.4 | 137 | 1.3 |  |
| Rather positive | 164 | 1.6 | 170 | 1.6 |  |
| Very positive | 27 | 0.3 | 36 | 0.3 |  |
| I do not know | 1502 | 14.4 | 1568 | 14.6 | 0.9 |

**eTable 3:** Impact of the Nutri-Score 2.0 on primary outcomes (i.e., objective understanding of nutritional quality and food ultra-processing) in stratified analyses, NutriNet-Santé, 2022, France, n = 21,159

|  | **OR* (95% CI) [experimental arm vs. control arm]** | | | ***P-trend*** | ***P-interaction*** |
| --- | --- | --- | --- | --- | --- |
| **In participants with a lower educational level (< high school)** | | | | |  |
| **All products** |  |  |  |  |  |
| **Number of correct answers** | 0 - 2 | 3 -6 | 7 - 9 |  |  |
| Understanding of nutritional quality | 1 | 0.81 (0.71 - 0.94) | 33.6 (15.7 - 71.8) | *<0.0001* | *<0.0001* |
| **Number of correct answers** | 0 - 12 | 13 - 19 | 20 - 22 |  |  |
| Understanding of ultra-processing | 1 | 2.06 (1.72 - 2.46) | 102.5 (71.9 - 146.2) | *<0.0001* | *<0.0001* |
| **In participants with a higher educational level (≥ high school)** | | | | |  |
| **All products** |  |  |  |  |  |
| **Number of correct answers** | 0 - 2 | 3 -6 | 7 - 9 |  |  |
| Understanding of nutritional quality | 1 | 0.49 (0.46 - 0.52) | 27.5 (22.0 - 34.4) | *<0.0001* | *<0.0001* |
| **Number of correct answers** | 0 - 12 | 13 - 19 | 20 - 22 |  |  |
| Understanding of ultra-processing | 1 | 1.91 (1.69 - 2.16) | 176.5 (151.0 - 206.4) | *<0.0001* | *<0.0001* |
| **In participants with a lower self-reported knowledge about nutrition** | | | | |  |
| **All products** |  |  |  |  |  |
| **Number of correct answers** | 0 - 2 | 3 -6 | 7 - 9 |  |  |
| Understanding of nutritional quality | 1 | 0.70 (0.62 - 0.80) | 60.7 (33.1 - 111.3) | *<0.0001* | *<0.0001* |
| **Number of correct answers** | 0 - 12 | 13 - 19 | 20 - 22 |  |  |
| Understanding of ultra-processing | 1 | 2.41 (1.97 - 2.93) | 341.3 (239.1 - 487.2) | *<0.0001* | *<0.0001* |
| **In participants with a higher self-reported knowledge about nutrition** | | | | |  |
| **All products** |  |  |  |  |  |
| **Number of correct answers** | 0 - 2 | 3 -6 | 7 - 9 |  |  |
| Understanding of nutritional quality | 1 | 0.50 (0.47 - 0.53) | 24.7 (19.6 - 31.1) | *<0.0001* | *<0.0001* |
| **Number of correct answers** | 0 - 12 | 13 - 19 | 20 - 22 |  |  |
| Understanding of ultra-processing | 1 | 1.82 (1.62 - 2.05) | 153.2 (131.0 – 179.2) | *<0.0001* | *<0.0001* |

**OR* = Odds Ratio**
ORs derived from multinomial logistic regression models to predict the number of correct answers according to the experimentation arm, adjusted for age, sex, educational level, household monthly income, professional situation and area of residence

**eTable 4:** Secondary outcomes: comparison of the 2 arms in terms of purchasing intentions and the healthiest-perceived product, NutriNet-Santé, 2022, n = 21,159

|  | **Control arm** | | **Experimental arm** | |  |
| --- | --- | --- | --- | --- | --- |
|  | **n** | **%** | **n** | **%** | ***p-value*** |
| **Look at these cookies** |  |  |  |  |  |
| **Which product would you most frequently buy?** |  |  |  |  |  |
| Cocoa and hazelnut stuffed cookies (Nutri-Score D, ultra-processed) | 421 | 4.1 | 233 | 2.2 |  |
| Butter shortbreads (Nutri-Score E, not ultra-processed) | 1371 | 13.2 | 982 | 9.1 |  |
| Granola chocolate chips cookies (Nutri-Score E, ultra-processed) | 808 | 7.8 | 423 | 3.9 |  |
| Chocolate chip oat bran cookies (Nutri-Score A, ultra-processed) | 380 | 3.7 | 707 | 6.6 |  |
| Double chocolate cookies (glazed and stuffed) (Nutri-Score E, ultra-processed) | 108 | 1.0 | 62 | 0.6 |  |
| Artisanal anise cookies (Nutri-Score C, not ultra-processed) | 1160 | 11.2 | 3678 | 34.2 |  |
| Oat bran whole organice cookies (Nutri-Score B, ultra-processed) | 2670 | 25.7 | 1300 | 12.1 |  |
| Organic hazelnut chocolate stuffed cookie (Nutri-Score D, not ultra-processed) | 579 | 5.6 | 1019 | 9.5 |  |
| None of the above | 2903 | 27.9 | 2355 | 21.9 | <0.0001 |
| **Which product seems the healthiest to you?** |  |  |  |  |  |
| Cocoa and hazelnut stuffed cookies (Nutri-Score D, ultra-processed) | 61 | 0.6 | 61 | 0.6 |  |
| Butter shortbreads (Nutri-Score E, not ultra-processed) | 593 | 5.7 | 240 | 2.2 |  |
| Granola chocolate chips cookies (Nutri-Score E, ultra-processed) | 42 | 0.4 | 50 | 0.5 |  |
| Chocolate chip oat bran cookies (Nutri-Score A, ultra-processed) | 1178 | 11.3 | 1527 | 14.2 |  |
| Double chocolate cookies (glazed and stuffed) (Nutri-Score E, ultra-processed) | 63 | 0.6 | 98 | 0.9 |  |
| Artisanal anise cookies (Nutri-Score C, not ultra-processed) | 2262 | 21.8 | 6664 | 61.9 |  |
| Oat bran whole organice cookies (Nutri-Score B, ultra-processed) | 4805 | 46.2 | 1069 | 9.9 |  |
| Organic hazelnut chocolate stuffed cookie (Nutri-Score D, not ultra-processed) | 328 | 3.2 | 332 | 3.1 |  |
| None of the above | 1068 | 10.3 | 718 | 6.7 | <0.0001 |
| **Do you think that the Nutri-Score 2.0 helped differentiate the nutritional quality of these cookies? (experimental arm)** |  |  |  |  |  |
| **Did you feel able to differentiate the nutritional quality of these cookies? (control arm)** |  |  |  |  |  |
| Yes, totally | 1052 | 10.1 | 3194 | 29.7 |  |
| Somehow, yes | 5074 | 48.8 | 5148 | 47.9 |  |
| No | 2727 | 26.2 | 1324 | 12.3 |  |
| Not at all | 1041 | 10.0 | 636 | 5.9 |  |
| I do not know | 506 | 4.9 | 457 | 4.3 | <0.0001 |
| **Do you think that the Nutri-Score 2.0 helped you identify among these cookies, those that are ultra-processed? (experimental arm)** |  |  |  |  |  |
| **Did you feel able to identify among these cookies, those that are ultra-processed? (control arm)** |  |  |  |  |  |
| Yes, totally | 931 | 9.0 | 7675 | 71.3 |  |
| Somehow, yes | 4477 | 43.1 | 2208 | 20.5 |  |
| No | 2694 | 25.9 | 338 | 3.1 |  |
| Not at all | 1698 | 16.3 | 281 | 2.6 |  |
| I do not know | 600 | 5.8 | 257 | 2.4 | <0.0001 |
| **Look at these breakfast cereals** |  |  |  |  |  |
| **Which product would you most frequently buy?** |  |  |  |  |  |
| Classical corn flakes (Nutri-Score D, ultra-processed) | 309 | 3.0 | 283 | 2.6 |  |
| Crispy chocolate muesli (Nutri-Score C, not ultra-processed) | 1027 | 9.9 | 1203 | 11.2 |  |
| Chocolate stuffed pillow-shaped cereals (Nutri-Score D, ultra-processed) | 205 | 2.0 | 198 | 1.8 |  |
| Chocolate flavoured corn flakes (Nutri-Score B, ultra-processed) | 271 | 2.6 | 356 | 3.3 |  |
| Sugar free fruit muesli (Nutri-Score A, not ultra-processed) | 3487 | 33.5 | 5964 | 55.4 |  |
| Chocolat oat crispy muesli (Nutri-Score D, ultra-processed) | 487 | 4.7 | 235 | 2.2 |  |
| Organic crispy muesli with red fruits (Nutri-Score D, not ulra-processed) | 956 | 9.2 | 345 | 3.2 |  |
| None of the above | 3658 | 35.2 | 2175 | 20.2 | <0.0001 |
| **Which product seems the healthiest to you?** |  |  |  |  |  |
| Classical corn flakes (Nutri-Score D, ultra-processed) | 233 | 2.2 | 311 | 2.9 |  |
| Crispy chocolate muesli (Nutri-Score C, not ultra-processed) | 819 | 7.9 | 460 | 4.3 |  |
| Chocolate stuffed pillow-shaped cereals (Nutri-Score D, ultra-processed) | 94 | 0.9 | 74 | 0.7 |  |
| Chocolate flavoured corn flakes (Nutri-Score B, ultra-processed) | 53 | 0.5 | 86 | 0.8 |  |
| Sugar free fruit muesli (Nutri-Score A, not ultra-processed) | 6515 | 62.6 | 9226 | 85.8 |  |
| Chocolat oat crispy muesli (Nutri-Score D, ultra-processed) | 72 | 0.7 | 38 | 0.4 |  |
| Organic crispy muesli with red fruits (Nutri-Score D, not ulra-processed) | 1339 | 12.9 | 147 | 1.4 |  |
| None of the above | 1275 | 12.3 | 417 | 3.9 | <0.0001 |
| **Do you think that the Nutri-Score 2.0 helped differentiate the nutritional quality of these cereals? (experimental arm)** |  |  |  |  |  |
| **Did you feel able to differentiate the nutritional quality of these cereals? (control arm)** |  |  |  |  |  |
| Yes, totally | 776 | 7.5 | 4827 | 44.9 |  |
| Somehow, yes | 4458 | 42.9 | 4283 | 39.8 |  |
| No | 2882 | 27.7 | 798 | 7.4 |  |
| Not at all | 1613 | 15.5 | 406 | 3.8 |  |
| I do not know | 671 | 6.5 | 445 | 4.1 | <0.0001 |
| **Do you think that the Nutri-Score 2.0 helped you identify among these cereals, those that are ultra-processed? (experimental arm)** |  |  |  |  |  |
| **Did you feel able to identify among these cereals, those that are ultra-processed? (control arm)** |  |  |  |  |  |
| Yes, totally | 894 | 8.6 | 7452 | 69.3 |  |
| Somehow, yes | 4301 | 41.4 | 2440 | 22.7 |  |
| No | 2785 | 26.8 | 337 | 3.1 |  |
| Not at all | 1783 | 17.1 | 234 | 2.2 |  |
| I do not know | 637 | 6.1 | 296 | 2.8 | <0.0001 |
| **Look at these ready-to-eat meals** |  |  |  |  |  |
| **Which product would you most frequently buy?** |  |  |  |  |  |
| Artisanal duck parmentier (Nutri-Score C, not ultra-processed) | 1324 | 12.7 | 1220 | 11.3 |  |
| Pasta box with bolognese sauce (Nutri-Score A, ultra-processed) | 164 | 1.6 | 437 | 4.1 |  |
| Pork sauté with mashed potatoes (Nutri-Score B, not ultra-processed) | 521 | 5.0 | 3429 | 31.9 |  |
| Gratinated endives with ham and emmental cheese (Nutri-Score B, ultra-processed) | 576 | 5.5 | 369 | 3.4 |  |
| Tartiflette (French traditional dish witth Reblochon cheese and oven cooked potatoes) (Nutri-Score C, ultra-processed) | 163 | 1.6 | 80 | 0.7 |  |
| Salmon with butter, shallots and mashed potatoes (Nutri-Score C, not ultra-processed) | 1533 | 14.7 | 1609 | 15.0 |  |
| Gratinated raviolis with scallops (Nutri-Score D, ultra-processed) | 776 | 7.5 | 230 | 2.1 |  |
| None of the above | 5343 | 51.4 | 3385 | 31.5 | <0.0001 |
| **Which product seems the healthiest to you?** |  |  |  |  |  |
| Artisanal duck parmentier (Nutri-Score C, not ultra-processed) | 1131 | 10.9 | 446 | 4.2 |  |
| Pasta box with bolognese sauce (Nutri-Score A, ultra-processed) | 183 | 1.8 | 899 | 8.4 |  |
| Pork sauté with mashed potatoes (Nutri-Score B, not ultra-processed) | 1323 | 12.7 | 7009 | 65.2 |  |
| Gratinated endives with ham and emmental cheese (Nutri-Score B, ultra-processed) | 1608 | 15.5 | 403 | 3.8 |  |
| Tartiflette (French traditional dish witth Reblochon cheese and oven cooked potatoes) (Nutri-Score C, ultra-processed) | 287 | 2.8 | 31 | 0.3 |  |
| Salmon with butter, shallots and mashed potatoes (Nutri-Score C, not ultra-processed) | 2650 | 25.5 | 1086 | 10.1 |  |
| Gratinated raviolis with scallops (Nutri-Score D, ultra-processed) | 563 | 5.4 | 48 | 0.5 |  |
| None of the above | 2655 | 25.5 | 837 | 7.8 | <0.0001 |
| **Do you think that the Nutri-Score 2.0 helped differentiate the nutritional quality of these meals? (experimental arm)** |  |  |  |  |  |
| **Did you feel able to differentiate the nutritional quality of these meals? (control arm)** |  |  |  |  |  |
| Yes, totally | 553 | 5.3 | 4322 | 40.2 |  |
| Somehow, yes | 3467 | 33.3 | 4327 | 40.2 |  |
| No | 3438 | 33.1 | 1083 | 10.1 |  |
| Not at all | 2191 | 21.1 | 513 | 4.8 |  |
| I do not know | 751 | 7.2 | 514 | 4.8 | <0.0001 |
| **Do you think that the Nutri-Score 2.0 helped you identify among these meals, those that are ultra-processed? (experimental arm)** |  |  |  |  |  |
| **Did you feel able to identify among these meals, those that are ultra-processed? (control arm)** |  |  |  |  |  |
| Yes, totally | 737 | 7.1 | 6968 | 64.8 |  |
| Somehow, yes | 3715 | 35.7 | 2743 | 25.5 |  |
| No | 3113 | 29.9 | 441 | 4.1 |  |
| Not at all | 2125 | 20.4 | 295 | 2.7 |  |
| I do not know | 710 | 6.8 | 312 | 2.9 | <0.0001 |

**eTable 5:** Perception of the Nutri-Score 2.0 by the experimental arm participants, NutriNet-Santé, 2022, n = 10,759

|  | **n** | **%** |
| --- | --- | --- |
| **The Nutri-Score 2.0 would help in my food purchases** |  |  |
| I do not agree at all | 400 | 3.7 |
| I somehow do not agree | 964 | 9.0 |
| I somhow agree | 5209 | 48.4 |
| I completely agree | 3833 | 35.6 |
| I do not know | 353 | 3.3 |
| **The Nutri-Score 2.0 gives me useful information for my food purchases** |  |  |
| I do not agree at all | 345 | 3.2 |
| I somehow do not agree | 924 | 8.6 |
| I somhow agree | 5005 | 46.5 |
| I completely agree | 4188 | 38.9 |
| I do not know | 297 | 2.8 |
| **The Nutri-Score is a misleading tool in food purchases** |  |  |
| I do not agree at all | 4377 | 40.7 |
| I somehow do not agree | 4175 | 38.8 |
| I somhow agree | 1241 | 11.5 |
| I completely agree | 408 | 3.8 |
| I do not know | 558 | 5.2 |
| **The Nutri-Score 2.0 is credible and trustworthy** |  |  |
| I do not agree at all | 289 | 2.7 |
| I somehow do not agree | 763 | 7.1 |
| I somhow agree | 5125 | 47.6 |
| I completely agree | 3353 | 31.2 |
| I do not know | 1229 | 11.4 |
| **The Nutri-Score 2.0 is easy to understand** |  |  |
| I do not agree at all | 265 | 2.5 |
| I somehow do not agree | 1018 | 9.5 |
| I somhow agree | 4961 | 46.1 |
| I completely agree | 4257 | 39.6 |
| I do not know | 258 | 2.4 |
| **I would like to see the Nutri-Score 2.0 displayed on food products** |  |  |
| I do not agree at all | 208 | 1.9 |
| I somehow do not agree | 342 | 3.2 |
| I somhow agree | 3205 | 29.8 |
| I completely agree | 6267 | 58.3 |
| I do not know | 737 | 6.9 |
| **The Nutri-Score 2.0 would be useful to inform me about the nutritional quality of food products** |  |  |
| I do not agree at all | 365 | 3.4 |
| I somehow do not agree | 970 | 9.0 |
| I somhow agree | 4698 | 43.7 |
| I completely agree | 4177 | 38.8 |
| I do not know | 549 | 5.1 |
| **The Nutri-Score 2.0 would be useful to help me identify ultra-processed foods** |  |  |
| I do not agree at all | 189 | 1.8 |
| I somehow do not agree | 241 | 2.2 |
| I somhow agree | 2172 | 20.2 |
| I completely agree | 7932 | 73.7 |
| I do not know | 225 | 2.1 |
| **I can rely on the Nutri-Score 2.0 to be informed on the nutritional quality of food products** |  |  |
| I do not agree at all | 401 | 3.7 |
| I somehow do not agree | 1093 | 10.2 |
| I somhow agree | 5029 | 46.7 |
| I completely agree | 3449 | 32.1 |
| I do not know | 787 | 7.3 |
| **I can rely on the Nutri-Score 2.0 to identify ultra-processed foods** |  |  |
| I do not agree at all | 208 | 1.9 |
| I somehow do not agree | 298 | 2.8 |
| I somhow agree | 2786 | 25.9 |
| I completely agree | 7113 | 66.1 |
| I do not know | 354 | 3.3 |
| **If I am not familiar with the product, I can rely on the Nutri-Score 2.0 to be informed on its nutritional quality** |  |  |
| I do not agree at all | 349 | 3.2 |
| I somehow do not agree | 1042 | 9.7 |
| I somhow agree | 5179 | 48.1 |
| I completely agree | 3586 | 33.3 |
| I do not know | 603 | 5.6 |
| **If I am not familiar with the product, I can rely on the Nutri-Score 2.0 to identify if it is ultra-processed** |  |  |
| I do not agree at all | 209 | 1.9 |
| I somehow do not agree | 262 | 2.4 |
| I somhow agree | 2759 | 25.6 |
| I completely agree | 7207 | 67.0 |
| I do not know | 322 | 3.0 |
